# The Australian Parkinson’s Genetics Study (APGS) - pilot (N = 1,532)

**DOI:** 10.1101/2021.04.02.21254825

**Authors:** Svetlana Bivol, George D. Mellick, Jacob Gratten, Richard Parker, Aoibhe Mulcahy, Philip Mosley, Peter C. Poortvliet, Adrian I. Campos, Brittany L. Mitchell, Luis M. Garcia-Marin, Simone Cross, Mary Ferguson, Penelope A. Lind, Danuta Z. Loesch, Peter M. Visscher, Sarah E. Medland, Clemens R. Scherzer, Nicholas G. Martin, Miguel E. Rentería

## Abstract

**Purpose:** Parkinson’s disease (PD) is a neurodegenerative disorder associated with progressive disability. While the precise aetiology is unknown, there is evidence of significant genetic and environmental influences on individual risk. The Australian Parkinson’s Genetics Study (APGS) seeks to study genetic and patient-reported data from a large cohort of individuals with PD in Australia to understand the sociodemographic, genetic, and environmental basis of PD susceptibility, symptoms and progression.

**Participants:** In the pilot phase reported here, 1,819 participants were recruited through assisted mailouts facilitated by *Services Australia* based on having three or more prescriptions for anti-PD medications in their Pharmaceutical Benefits Scheme (PBS) records. The average age at the time of the questionnaire was 64 ± 6 years. We collected patient-reported PD information and socio-demographic variables via an online (93% of the cohort) or paper-based (7%) questionnaire. One thousand five hundred thirty-two participants (84.2%) met all inclusion criteria, and 1,499 provided a DNA sample via traditional post.

**Findings to date:** 65% of participants were male, and 92% identified as being of European descent. A previous traumatic brain injury was reported by 16% of participants and was correlated with a younger age of symptom onset. At the time of the questionnaire, constipation (36% of participants), depression (34%), anxiety (17%), melanoma (16%) and diabetes (10%) were the most reported comorbid conditions.

**Future plans:** We plan to recruit sex- and age-matched unaffected controls, genotype all participants, and collect non-motor symptoms and cognitive function data. Future work will explore the role of genetic and environmental factors in the aetiology of PD susceptibility, onset, symptoms, and progression, including as part of international PD research consortia.

**Article summary:** *Strengths and limitations of this study:*

- We used a time- and cost-effective recruitment method that enabled us to reach out to a random sample of individuals who have been prescribed medications for Parkinson’s disease across all over Australia to invite them to participate in this study.
- The identities of letter recipients remained private and confidential and were not shared with the researchers. However, those recipients who were interested and willing to participate were directed to a website where they could sign up and provide informed consent.
- The source database only captures individuals who have been prescribed medications to treat Parkinson’s disease in Australia and who are eligible for Medicare. Those without an official diagnosis, not receiving treatment, or not eligible for government subsidies are not included.
- We collected a wide range of patient-reported variables relevant to disease onset, diagnosis, symptoms, medical comorbidities, lifestyle, and family history in a large cohort of participants. However, some variables might not be as accurate as when measured by a specialist clinician.
- Given the 9% response rate to our single-letter invitation, there is a substantial risk of self-selection bias. Thus, patient characteristics for this cohort might differ from those of the typical population of individuals with Parkinson’s disease in Australia.

## Introduction

Parkinson’s disease (PD) is a progressive neurodegenerative disorder that affects 2-3% of the global population over 65 years [1,2]. At present, over 100,000 Australians have PD, and approximately 32 new cases are diagnosed every day [3]. The economic cost of Parkinson’s disease in Australia was estimated at almost $10 billion per year in 2015, but the prevalence and total financial cost associated with the disease have increased dramatically since then [4]. The number of newly diagnosed PD cases is expected to double over the coming decades due to population ageing, posing substantial challenges to the healthcare system, the economy, and society [5].

The pathological hallmarks of PD include nigrostriatal degeneration and widespread intracellular accumulation of Lewy bodies in multiple brain regions. The aetiological pathways of PD are mostly unknown. Many risk factors have been explored, including certain drugs, head trauma, lifestyle factors, comorbid diseases, and exposure to toxic environmental agents [6–8]. In particular, age, mood disorders, consumption of well water [9], head injury [10], and exposure to environmental toxins [11,12] have been consistently associated with an increased PD risk. On the contrary, smoking and caffeine have been inversely related to PD risk and positively correlated with a later age of onset [13,14], suggesting a putative protective effect that needs further exploration. Associations with alcohol remain controversial [15].

The clinical manifestations of PD are highly heterogeneous. Diagnosis is based on the presence of cardinal motor symptoms, including bradykinesia, resting tremor, muscle rigidity, gait difficulty, and postural instability [1]. Non-motor symptoms are also common, particularly in late-stage PD, and involve cognitive impairment (e.g., executive dysfunction and attentional deficits), psychotic symptoms (e.g., visual hallucinations), autonomic dysfunction (e.g., orthostatic hypotension, urogenital incontinence, and constipation), disorders of sleep and mood, hyposmia, fatigue and chronic pain [16]. Notably, some comorbid medical conditions such as constipation, hyposmia, REM sleep behaviour disorder, diabetes and mood disorders may precede the onset of motor disability by years or even decades and serve as pre-diagnostic PD symptoms [17]. Thus, it is worth noting that PD onset, clinical presentation, and progression differ substantially among patients [18].

PD has a complex genetic architecture, with both monogenic and sporadic forms. A twin study in the Swedish Twin Registry estimated heritability at 34%, with concordance rates for PD of 11% for monozygotic twins and 4% for same-sexed dizygotic twin pairs [19]. Rare coding variants detected in the *LRRK2, VPS12C, GBA* and *SNCA* genes are associated with monogenic (familial) PD forms [20], whereas common variants around the same *GBA, SNCA* and *LRRK2* genes have also been associated with sporadic (non-familial) PD [21]. A recent GWAS meta-analysis of 17 cohorts identified 90 independent single-nucleotide polymorphisms (SNPs) across 78 genomic regions associated with sporadic Parkinson’s risk [22]. Despite this remarkable progress, common genetic variants only explain around 22% (95% CI 18% - 26%) of PD risk on the liability scale, implying that there are still many more unidentified genetic variants contributing to PD susceptibility. It has been pointed out that the inclusion of diverse populations in genetic studies improves the utility of genetic discovery [23]. However, genetic PD studies have focused mainly on European ancestry individuals, limiting their capacity to extrapolate results to other ethnicities. Australia is ethnically diverse, but the disease epidemiology among various ethnic groups has not been investigated [24,25].

Treatment response in PD is complex and poorly understood [26]. Although there is currently no cure for PD, a combination of pharmacological (e.g., levodopa, dopamine receptor agonists, anticholinergics) and non-pharmacological (e.g., physiotherapy, deep brain stimulation) therapies are the only available treatment options. However, such interventions only ameliorate the symptoms of the illness, and their long term use is associated with adverse side effects in some patients [27,28], which in turn result in suboptimal medication compliance and low quality of life. Research seeking to identify the disease’s genetic components that contribute to drug response variability could lead to the discovery of personalised disease-modifying therapies with great potential to improve PD patients’ well-being and quality of life.

Few studies have assessed the prevalence, risk factors, sociodemographic and genetic factors of PD in Australia. Similarly, details about disease onset, symptoms and progression and other medical comorbidities have not been collected and studied at a large scale in a nationwide sample. Long-running studies, such as the Griffith University Queensland Parkinson’s Project, have made significant contributions over the past 20 years in describing risk factors and mechanisms underlying PD’s pathological development and its disease progression [29–31]. Despite continued global scientific efforts, many questions remain around early and accurate diagnosis, differentiation, prognostication, better traditional management methods and new treatment efforts such as neuroprotection and disease modification. With an aging population, the number of individuals with PD in Australia is set to increase dramatically. Thus, it is crucial to generate reliable evidence about the epidemiology and genetic aetiology of PD to inform policy and clinical practice. Ultimately, a more extensive representation of diverse Australian participants in worldwide PD studies may also lead to discovering novel therapeutic targets and developing new therapies and interventions to prevent, stop, or modify PD’s clinical course in Australia and the rest of the world.

### Hypothesis

We hypothesise that individual differences in PD susceptibility, symptomatology, progression, and treatment response result, in part, from underlying individual genetic variability. Thus, large longitudinal cohorts of individuals with PD will be required to unravel the complex aetiology of PD heterogeneity.

### Aim of the study

The Australian Parkinson’s Genetics Study aims to recruit and follow-up a large cohort of Australians with PD and unaffected controls and characterise their phenotypic, genetic, and environmental diversity. Here we describe the results of a pilot study comprising the first ∼1,500 participants.

### Objectives

1. To recruit and follow up a nationwide cohort of 10,000 participants with PD and 10,000 unaffected controls and characterise their genetic information.
2. To collect patient-reported data on disease onset, symptoms, comorbidities, and progression; treatment response; environmental, lifestyle and sociodemographic factors; family and medical history; and cognitive function.
3. Discover and validate genetic and environmental markers of PD susceptibility and heterogeneity and contribute to identifying and validating therapeutic molecular targets.
4. To contribute to ongoing international collaborative PD research efforts.

## Methodology

The present study used a cross-sectional study design. Here we describe the design of the Australian Parkinson’s Genetics pilot study and baseline characteristics of participants, including data collection methods, sociodemographic variables, environmental and lifestyle exposures, and self-reported history of medical conditions. Participants in the present study were Australian residents diagnosed with PD. The pilot project was initially conceived by researchers at the QIMR Berghofer Medical Research Institute (QIMRB) and approved by QIMRB’s Human Research Ethics Committee, with all participants providing informed consent for participation in the study and DNA genotyping.

### Recruitment method

Participants were invited to participate in the study via assisted mailouts through *Services Australia* (formerly known as the Australian Government Department of Human Services) based on their pharmaceutical prescription history. *Services Australia* is an official government agency responsible for delivering a range of welfare, health, child support payments and other services to eligible Australian citizens and permanent residents, including managing the Pharmaceutical Benefits Scheme (PBS). The PBS is a program of the Australian Government that subsidises prescription medication. Under the PBS, all Australian residents holding a current Medicare card are eligible for prescription drug subsidies to make commonly used medicines more accessible and affordable (for detailed information about the PBS and a list of all medicines on the PBS, refer to the following website https://www.pbs.gov.au/pbs/home). *Services Australia* retains records for the most recent five years’ PBS-subsidized medicines and considers requests for assisted mailouts from external organisations for health-related research projects. External organisations must have approval from a Human Research Ethics Committee, per the National Health and Medical Research Council’s guidelines. *Services Australia* then sends the approach letters to individuals meeting specific selection criteria on behalf of the research organisation. Personal data of letters recipients is not shared with the researchers requesting the mailout and thus always remains secure and confidential.

In this pilot phase, 20,000 invitation letters were sent in June 2020 to individuals who: (1) were Australian residents (Australian citizens or permanent residents), aged 40-75 years; (2) had at least three PBS claims in the last two years for medications commonly used in the management of symptoms of Parkinson’s disease (see **Supplementary Table 1** for a list of medications and their corresponding PBS codes) according to their records in the PBS database. The invitation letters directed potential participants to a secure study website (https://www.qimrberghofer.edu.au/parkinsonsgenetics) with more detailed information about the study. Individuals who did not have PD were kindly asked to ignore the invitation letter. Those interested to participate were given the option to complete an online or a paper-based questionnaire. Participants indicated having a current clinical diagnosis for PD and the type of clinician who initially diagnosed them (e.g., neurologist, geriatrician, general practitioner). Prospective participants were also asked to indicate their consent to donate a saliva sample for DNA extraction and genotyping.

Those willing to provide a saliva sample were sent a saliva DNA collection kit via mail. A small percentage of prospective participants (∼ 10%) requested to be mailed a paper-based version of the questionnaire and consent form. Of the 1,819 individuals who filled out the registration form and provided informed consent to participate (9.1% response rate for the mailouts), 1,532 (84%) confirmed having a current PD diagnosis by a licensed clinician and were willing to provide a DNA sample.

Of those who consented to donate their saliva for genetic analysis, 1,499 (97.8%) have returned saliva kits by mail as of August 2021. Since information about the letter recipients remained confidential, we could not follow up with the nonrespondents, some of whom may have died or moved to a new address. Refer to **Supplementary Table 2** for the percentage of APGS invitations sent and participant registration per Australian state or territory.

We are aware of a few cases where a letter recipient shared the invitation with other relatives or acquaintances with PD who did not initially receive an approach letter. This became evident after a few participants aged over 75 (the upper limit of our target sample for the mailout) registered in the study. Unfortunately, we cannot reliably quantify the number of participants who signed up but did not receive an invitation letter from *Services Australia*. None of the participants was excluded based on age if they met the other inclusion criteria of the study.

### Patient and Public Involvement

During the project, we reached out to patient support groups and advocacy organisations. Notably, the Shake It Up Australia Foundation for Parkinson’s Research (www.shakeitup.org.au) has become an ally of the project. When the full-fledged study is rolled out, a patient advisory group will be formed to provide input and feedback about the research agenda. Similarly, the research team will share research updates at least once yearly with the participants via an electronic or physical newsletter.

### Genotyping plans

Genotyping will be conducted using the Illumina Global Screening Array. Genotypes will be processed and subjected to quality control, and genotype imputation will be conducted using state of the art protocols. We will use available genetic controls from screened population-based studies at QIMR to perform several analyses, including estimating SNP-based heritability with GCTA, a GWAS meta-analysis to combine our results with the summary results from previous GWASs of PD, or estimating polygenic risk scores for relevant comorbid conditions. We plan to recruit demographically matched controls in subsequent waves of the study.

### Data collection

The online instruments consisted of several sections, covering sociodemographic, clinical, occupational, lifestyle and environmental factors, with a schematic of the APGS and questionnaire content shown in **Figure 1**. As part of the informed consent process, all participants agreed to be recontacted for future related studies, understanding that their participation in future studies is voluntary. The second wave of data collection is planned for the first half of 2022. It will include questions assessing heterogeneity in PD onset and motor and non-motor disability, including the following instruments: items from the Movement Disorder Society United Parkinson’s Disease Rating Scale (MDS-UPDRS) [32]; a short scale for the assessment of motor impairments and disability in Parkinson’s disease (The SPES/SCOPA); the Apathy Screening Tool; Geriatric Anxiety Inventory [32,33]; Geriatric Depression Scale (GDS) short-form [34]; Assessment of Psychiatric Complications in Parkinson’s disease (The SCOPA-PC) [35]; and the SCOPA-sleep and PDSS scales [36] to identify sleep disorders in PD.

**Figure 1.**
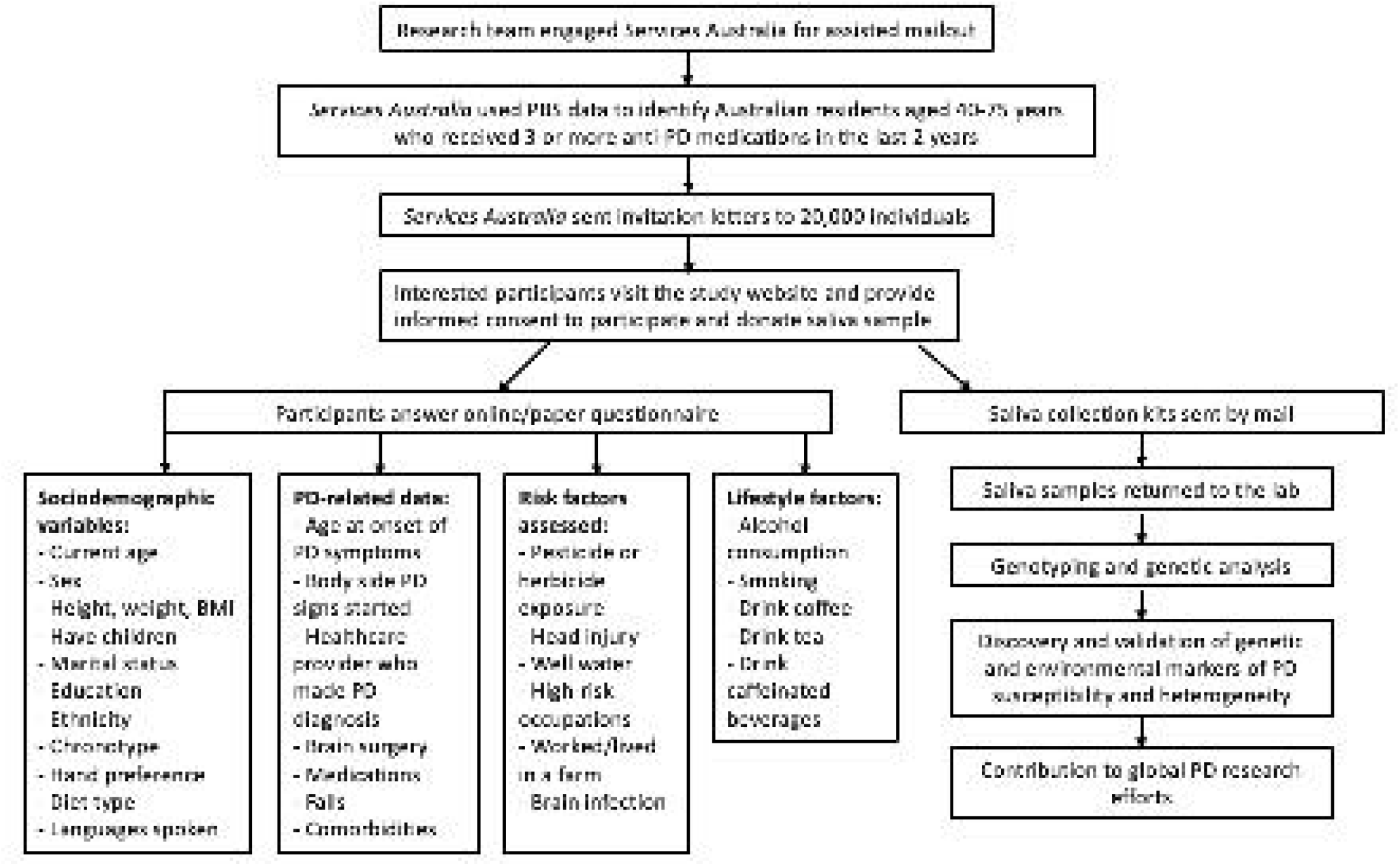
Overview of the recruitment strategy and questionnaire contents for the APGS pilot.

### Statistical analyses

IBM SPSS Statistics (Version 23) software was used for statistical analysis [37]. In between-group analysis (male versus female), *P*-values were calculated using a t-test for comparing mean values of the samples or a Chi-square test for comparing frequencies. We used a bivariate correlation test (e.g., Pearson correlation coefficient) to assess a relationship between two continuous variables. We used an alpha level of 0.05 for all statistical tests.

## Results

We recruited 1,532 participants (mean age = 64 ± 6 years; see **Supplementary Figure 1** and **Supplementary Figure 2** for sex–stratified age distributions). Sociodemographic data were collected via web-based or paper questionnaires, with most participants (93%; N = 1,424) responding via the online questionnaire. Of our sample, 65% were males, and the mean BMI of the sample was 28 ± 5kg/m^2^.

Most participants reported being diagnosed with PD by a neurologist (83%), having children (87%), being married or in a de facto relationship (81%) and being of European ancestry (92%). Nearly 60% have a post-high school qualification, and 15% speak more than one language. Tremor was the most common first symptom of PD reported in 54% of the sample, followed by bradykinesia and muscle rigidity (20%), followed by gait disorder (10%), with an average age at onset of the first PD symptom of 57 ± 8 years. The frequency of the first self-reported symptom of PD in participants is presented in **Figure 2**.

**Figure 2.**
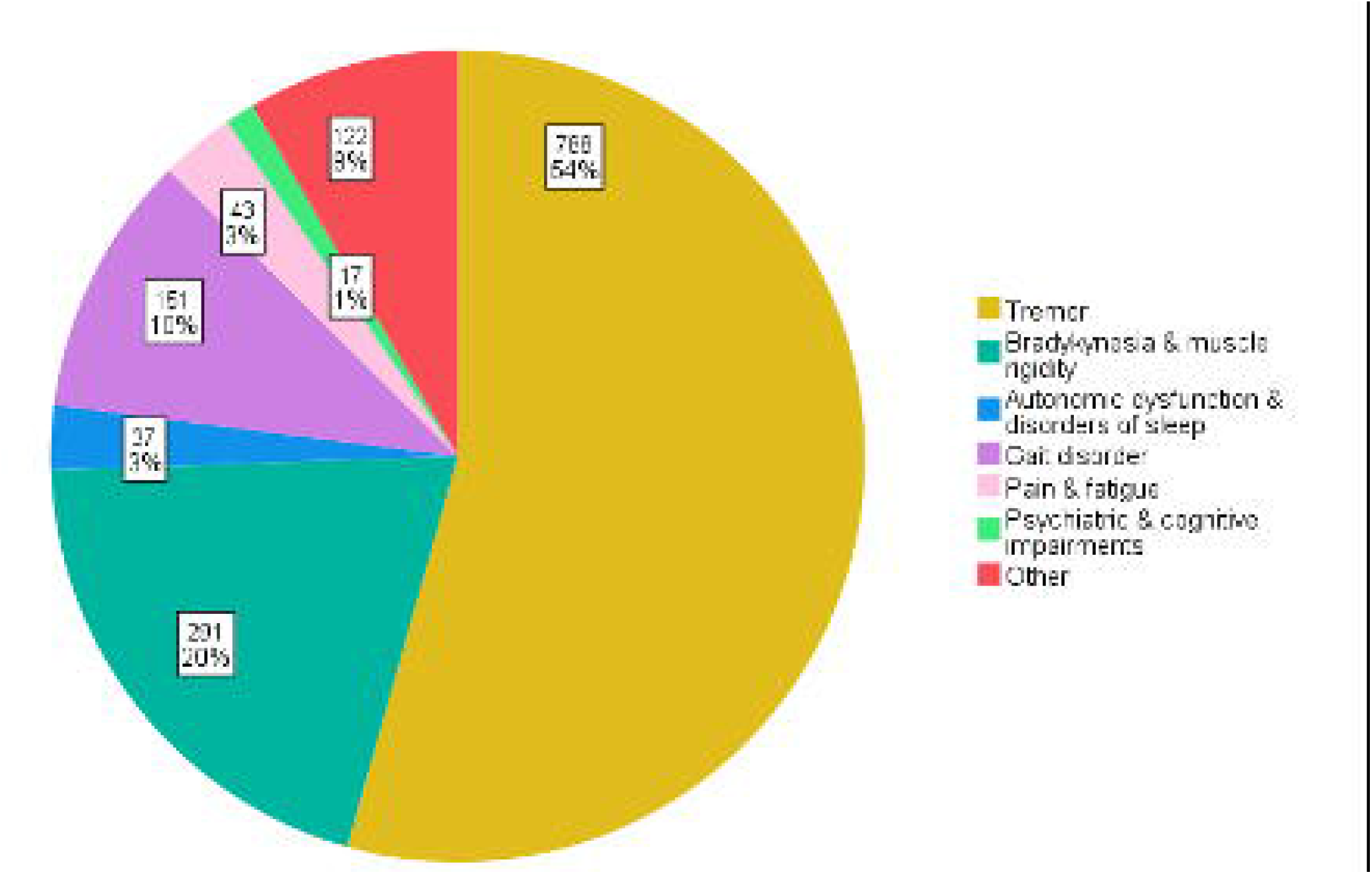
Frequency of patient-reported first symptom of participants in the APGS pilot cohort.

For 85% of participants, PD symptoms started on one side of the body, and more than half (44%) experienced the symptoms on the right side of the body. Most respondents (94%) were prescribed at least one anti-PD medication at the time of the questionnaire. Patients who were not on anti-PD medication were included in the study only if they had received a DBS surgery. Furthermore, 42% had experienced PD-related falls, and 12% reported having had brain surgery for PD. The frequencies and percentages of all self-reported sociodemographic and PD-related characteristics are outlined in **Table 1** and **Table 2**, respectively.

**Table 1.** Sociodemographic characteristics of study participants

|  | **Total**<br>**N (%)** | **Male**<br>**N (%)** | **Female**<br>**N (%)** | **P-values Male vs**<br>**Female** |
| --- | --- | --- | --- | --- |
| Response method |  |  |  | 0.87 |
| On-line questionnaire | 1424 (92.9) | 924 (92.8) | 499 (93.1) |  |
| Paper questionnaire | 108 (7.1) | 71 (7.2) | 37 (6.9) |  |
| Age in years |  |  |  |  |
| Mean (SD) | 63.9 (5.9) | 64.2 (5.9) | 63.5 (6.0) | 0.04 |
| Range | 40 – 88 | 40 - 86 | 42 - 88 |  |
| Sex |  |  |  |  |
| Female | 536 (35.0) |  |  | - |
| Male | 995 (64.9) |  |  | - |
| MTF | 1 (0.06) |  |  | - |
| Current height in cm |  |  |  |  |
| Mean (SD) | 170.7 (9.9) | 175.4 (7.8) | 162.0 (7.2) | <0.001 |
| Range | 124 - 200 | 150 - 200 | 124 - 178 |  |
| Current weight in kg |  |  |  |  |
| Mean (SD) | 80.8 (16.9) | 85.5 (15.3) | 72.3 (16.5) | <0.001 |
| Range | 40 - 164 | 45 - 164 | 40 - 140 |  |
| Current BMI (kg/m<sup>2</sup>) |  |  |  |  |
| Mean (SD) | 27.6 (5.3) | 27.7 (4.8) | 27.5 (6.0) | 0.29 |
| Range | 16.6 – 55.1 | 17.5 – 50.7 | 16.6 – 55.1 |  |
| Have children | 1338 (87.3) | 870 (84.4) | 467 (87.1) | 0.86 |
| Marital status |  |  |  | <0.001 |
| Single / never married | 66 (4.3) | 45 (4.5) | 21 (3.9) |  |
| Married / de facto relationship | 1245 (81.3) | 848 (85.3) | 396 (74.0) |  |
| Separated / divorced | 145 (9.5) | 70 (7.0) | 75 (14.0) |  |
| Widowed | 48 (3.1) | 17 (1.7) | 31 (5.7) |  |
| In a relationship but not living<br>together | 26 (1.7) | 13 (1.3) | 13 (2.4) |  |
| Other | 2 (0.1) | 2 (0.2) | 0 (0.0) |  |
| Ethnicity |  |  |  | 0.73 |
| European | 1397 (91.6) | 910 (91.5) | 486 (90.7) |  |
| East Asian | 40 (2.6) | 23 (2.4) | 17 (3.2) |  |
| Indigenous Australian / Torres Strait<br>Islander | 12 (0.8) | 8 (0.7) | 4 (0.7) |  |
| African | 2 (0.1) | 2 (0.2) | 0 (0.0) |  |
| South Asian | 26 (1.7) | 14 (1.4) | 12 (2.2) |  |
| Pacific Islander | 9 (0.6) | 6 (0.6) | 3 (0.6) |  |
| Hispanics | 5 (0.3) | 4 (0.4) | 1 (0.2) |  |
| Mixed | 21 (1.4) | 16 (1.7) | 5 (0.9) |  |
| Other | 13 (0.8) | 8 (0.7) | 5 (0.9) |  |
| Information not provided | 7 (0.4) | 4 (0.4) | 3 (0.6) |  |
| Education |  |  |  | 0.02 |
| Junior high school or less | 348 (22.7) | 218 (21.9) | 130 (24.3) |  |
| Senior high school | 276 (18.0) | 184 (18.5) | 92 (17.3) |  |
| Certificate III/IV | 196 (12.8) | 124 (12.5) | 72 (13.4) |  |
| Diploma | 190 (12.4) | 128 (12.8) | 62 (11.6) |  |
| Bachelor's degree | 235 (15.3) | 163 (16.4) | 71 (13.3) |  |
| Graduate diploma or certificate | 134 (8.7) | 77 (7.7) | 57 (10.6) |  |
| Postgraduate | 148 (9.7) | 97 (9.7) | 51 (9.5) |  |
| Information not provided | 5 (0.3) | 5 (0.5) | 0 (0.0) |  |
| Hand preference |  |  |  | 0.78 |
| Left hand | 156 (10.2) | 103 (10.4) | 53 (9.9) |  |
| Right hand | 1323 (86.5) | 856 (86.1) | 467 (87.3) |  |
| Both hands | 50 (3.3) | 34 (3.4) | 15 (2.8) |  |
| Not sure | 1 (0.06) | 1 (0.1) | 0 (0.0) |  |
| Chronotype |  |  |  | <0.001 |
| Morning-type person | 800 (52.3) | 518 (52.2) | 282 (52.6) |  |
| Evening-type person | 376 (24.6) | 212 (21.3) | 163 (30.4) |  |
| Neither | 355 (23.2) | 264 (26.5) | 91 (17.0) |  |
| Information not provided | 1 (0.06) | 1 (0.1) | 0 (0.0) |  |
| Consider themselves spiritual or religious | 599 (39.1) | 347 (34.9) | 251 (46.8) | <0.001 |
| Information not provided | 3 (0.2) | 2 (0.2) | 1 (0.2) |  |
| Diet type |  |  |  | 0.004 |
| Vegan | 8 (0.5) | 7 (0.7) | 1 (0.2) |  |
| Vegetarian | 42 (2.7) | 18 (1.8) | 22 (4.1) |  |
| Speak more than one language | 228 (14.9) | 147 (14.8) | 81 (15.1) | 0.88 |
| Information not provided | 1 (0.06) | 1 (0.1) | 0 (0.0) |  |

**Note:** MTF – male-to-female transgender person; SD – standard deviation; BMI – body mass index; *P*-values were calculated using a t-test for comparing mean values or a Chi-square test for comparing frequencies; Sample N=1532.

**Table 2.** Parkinson’s disease specific outcomes

|  | Total<br>N (%) | Male<br>N (%) | Female<br>N (%) | *P*-values Male vs<br>Female |
| --- | --- | --- | --- | --- |
| Age at onset of the first PD symptom |  |  |  |  |
| Mean (SD) | 56.1 (8.3) | 56.4 (8.2) | 55.6 (8.3) | 0.10 |
| Range | 27 - 86 | 30 - 77 | 27 - 86 |  |
| Healthcare provider who made PD diagnosis |  |  |  | 0.001 |
| Neurologist | 1260 (82.2) | 790 (79.4) | 469 (87.5) |  |
| Geriatrician | 16 (1.0) | 12 (1.2) | 4 (0.7) |  |
| General practitioner | 237 (15.5) | 178 (17.9) | 59 (11.0) |  |
| Other | 15 (0.9) | 12 (1.2) | 3 (0.6) |  |
| Information not provided | 4 (0.3) | 3 (0.3) | 1 (0.2) |  |
| Had brain surgery for PD | 177 (11.5) | 119 (11.9) | 58 (10.8) | 0.53 |
| Take at least one medication for PD | 1424 (93.9) | 930 (93.5) | 493 (92.0) | 0.43 |
| Experienced falls related to PD | 631 (41.2) | 383 (38.5) | 247 (46.1) | 0.004 |
| Information not provided | 13 (0.8) | 8 (0.8) | 5 (0.9) |  |
| PD signs started on one side of the body | 1296 (84.6) | 833 (83.6) | 463 (86.3) | 0.12 |
| Information not provided | 9 (0.6) | 6 (0.6) | 3 (0.5) |  |
| Side of the body where PD symptoms started |  |  |  | 0.69 |
| Left | 625 (40.8) | 405 (40.7) | 220 (41.0) |  |
| Right | 671 (43.8) | 427 (42.9) | 243 (45.3) |  |

### Occupational and environmental risk factors of PD

Previously reported occupational (e.g., welding, metal melting, agriculture, petrochemistry) and environmental risk factors associated with PD were examined in this cohort and included exposure to pesticides or herbicides, head injury, well water drinking, working in high-risk occupations, living on a farm for over a year, and having metal poisoning or brain infection at any point across the life course. Thirty-four per cent of the sample reported being exposed to pesticides or herbicides at some point in their lives. Of those exposed to pesticides/herbicides, 390 were male, and 127 were female, with most of these cases (82%) reporting pesticide/herbicide exposure before 37 years of age.

Sixteen per cent of the sample had experienced head trauma, and the average age of head injury was 26 ± 18 years. Of those with a previous head injury, 67% reported traumatic head injury with loss of consciousness. There was a significant positive association between the age of traumatic head injury and the age at onset of the first symptom of PD (Pearson correlation coefficient r = 0.3, *p* < 0.001) (**Figure 3**).

**Figure 3.**
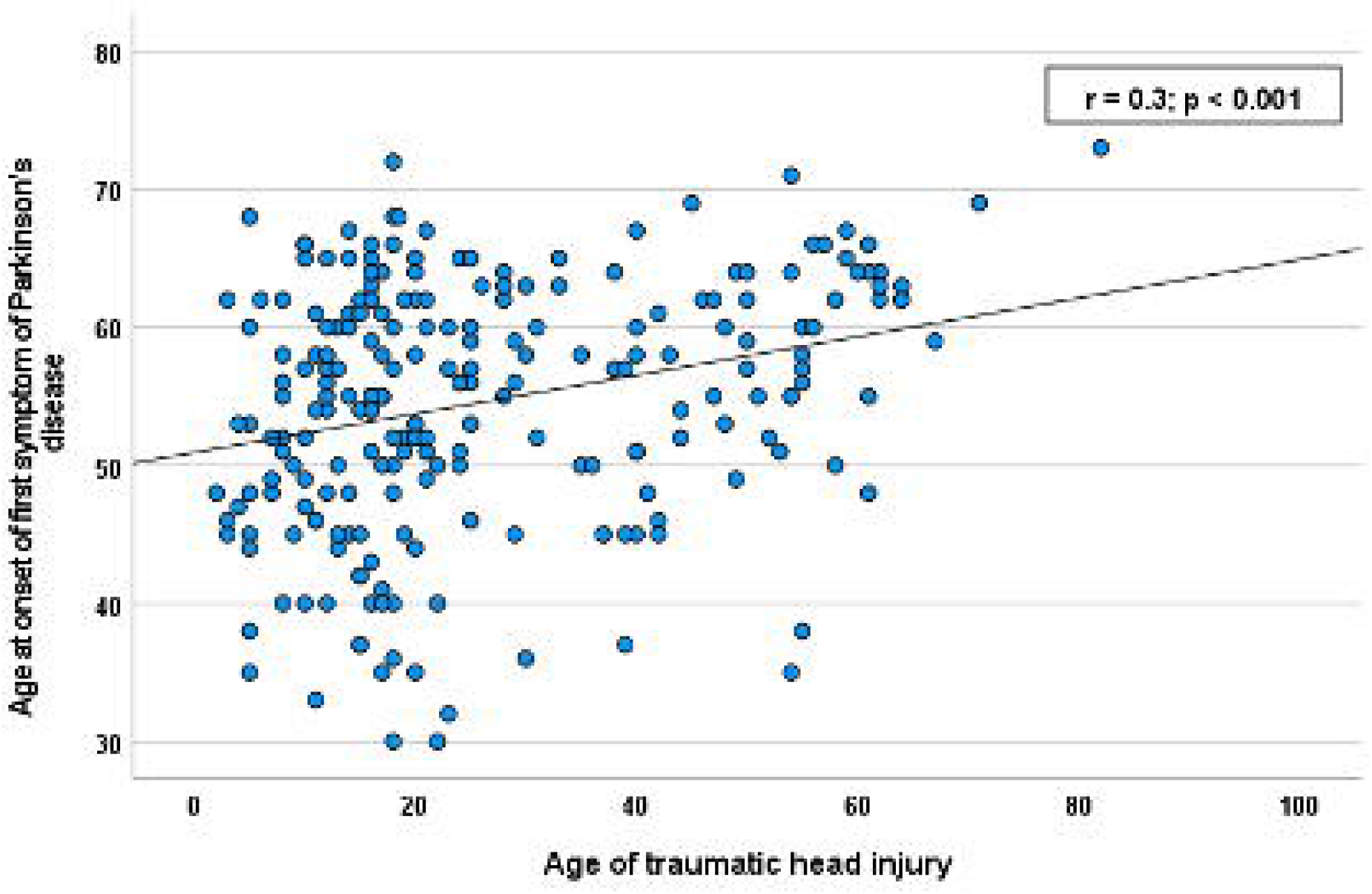
Traumatic brain injury is correlated with age at onset of first PD symptom.

Five per cent of participants reported having frequently consumed water from a well for 19 ± 12 years on average. A third of the sample (33%) disclosed having worked in a high-risk occupation throughout their lives, including welding, metal melting, galvanisation, milling, petrochemistry, agriculture, wood processing, and textile or industrial painting. Of these participants, 42% reported exposure to more than one occupational risk factor. Approximately 24% of the cohort reported having worked or lived on a farm for more than a year, 1% acquired metal poisoning and 2% encephalitis (brain infection). The occupational and environmental risk factors of PD, summarised by sex, are shown in **Table 3**.

**Table 3.** Occupational and environmental risk factors

| Risk factor | Total<br>N (%) | Male<br>N (%) | Female<br>N (%) |
| --- | --- | --- | --- |
| Exposure to pesticides or herbicides | 517 (33.6) | 390 (39.2) | 127 (23.6) |
| Information not provided | 25 (1.6) | 16 (1.6) | 9 (1.7) |
| Age of exposure to pesticides or herbicides |  |  |  |
| < 18 years of age | 185 (12.1) | 128 (12.9) | 57 (10.6) |
| 18 – 36 | 239 (15.6) | 190 (19.1) | 49 (9.1) |
| 37 – 55 | 84 (5.5) | 64 (6.4) | 20 (3.7) |
| ≥ 56 | 9 (0.6) | 8 (0.8) | 1 (0.2) |
| Head injury | 244 (15.9) | 186 (18.7) | 58 (10.8) |
| Information not provided | 21 (1.4) | 12 (1.2) | 9 (1.7) |
| Age of head injury |  |  |  |
| Mean (SD) | 26.3 (17.6) | 26.2 (16.5) | 26.9 (20.8) |
| Range | 2 - 82 | 3 - 67 | 2 - 82 |
| Head injury with loss of consciousness | 165 (10.8) | 128 (12.8) | 37 (6.9) |
| Well water drinking | 82 (5.3) | 55 (5.5) | 27 (5.0) |
| Information not provided | 34 (2.2) | 21 (2.1) | 13 (2.4) |
| Number of years drinking well water |  |  |  |
| Mean (SD) | 18.7 (12.4) | 18.3 (12.9) | 19.4 (11.6) |
| Range | 2 - 50 | 2 - 50 | 4 - 40 |
| Worked in high-risk occupations | 504 (32.9) | 425 (42.7) | 79 (14.7) |
| Number of high-risk occupations worked throughout of life |  |  |  |
| One | 293 (19.1) | 226 (22.8) | 67 (12.5) |
| Two | 115 (7.5) | 107 (10.7) | 8 (1.5) |
| Three | 52 (3.4) | 48 (4.8) | 4 (0.7) |
| Four | 29 (1.9) | 29 (2.9) | 0 (0.0) |
| Five or more | 15 (0.9) | 15 (1.5) | 0 (0.0) |
| Worked or lived on a farm for over 1 year | 360 (23.5) | 236 (23.7) | 124 (23.1) |
| Information not provided | 27 (1.8) | 18 (1.8) | 9 (1.6) |
| Had metal poisoning | 22 (1.4) | 17 (1.7) | 5 (0.9) |
| Information not provided | 34 (2.2) | 21 (2.1) | 13 (2.4) |
| Had brain infection (encephalitis) | 31 (2.0) | 20 (2.0) | 11 (2.1) |
| Information not provided | 21 (1.4) | 12 (1.2) | 1 (0.2) |

**Note:** High-risk occupations included welding, metal melting, galvanization, milling, petrochemistry, agriculture, wood processing, textile, or industrial painting; SD – standard deviation; N = 1532

### Lifestyle factors correlated with lower PD risk

As shown in **Table 4**, we investigated a range of lifestyle variables that have been correlated in previous PD studies with lower PD risk. These included alcohol drinking, smoking, exposure to second-hand smoke, and drinking caffeinated coffee, tea, and other beverages. More than two-thirds of participants (71%) currently drink an average of 8 ±8 standard alcoholic drinks per week, and 20% reported consuming alcohol heavily in the past. There was a significant weak positive association between the number of alcoholic drinks consumed per week and the age at onset of the first PD symptom (Pearson correlation coefficient r = 0.1, *p* =0.001).

**Table 4.** Lifestyle variables

|  | **Total**<br>**N (%)** | **Male**<br>**N (%)** | **Female**<br>**N (%)** |
| --- | --- | --- | --- |
| Currently drink alcohol | 1090 (71.1) | 751 (75.5) | 339 (63.2) |
| Mean number of drinks per week (SD) | 7.6 (7.7) | 8.3 (8.4) | 5.7 (5.3) |
| Range | 1 - 50 | 1 - 50 | 1 - 30 |
| Information not provided | 37 (2.4) | 23 (2.3) | 14 (2.6) |
| Consumed alcohol heavily in the past | 313 (20.4) | 252 (25.3) | 61 (11.4) |
| Current cigarette/cigar/pipe smokers | 50 (3.3) | 28 (2.8) | 22 (4.1) |
| Mean number of cigarettes/cigar/pipes smoked per day (SD) | 13.3 (8.3) | 14.3 (8.9) | 12.0 (7.6) |
| Range | 2 - 40 | 2 - 40 | 2 - 30 |
| Former smokers | 554 (36.2) | 368 (37.0) | 186 (34.7) |
| Mean number of cigarettes smoked per day (SD) | 15.3 (11.2) | 18.1 (4.6) | 17.9 (3.5) |
| Range | 1 - 60 | 10 - 49 | 10 - 31 |
| Information not provided | 37 (2.4) | 24 (2.4) | 13 (2.4) |
| Exposure to second-hand smoke at home | 858 (56.0) | 510 (51.3) | 348 (64.9) |
| Mean number of hours per day (SD) | 4.5 (3.7) | 4.3 (3.6) | 4.9 (3.9) |
| Range | 0.5 - 20 | 0.5 - 18 | 0.5 - 20 |
| Exposure to second-hand smoke at work | 701 (45.7) | 521 (52.4) | 179 (33.4) |
| Mean number of hours per day (SD) | 4.9 (3.1) | 4.9 (3.2) | 4.9 (2.9) |
| Range | 0.5 - 12 | 0.5 - 12 | 0.5 - 12 |
| Exposure to second-hand smoke in other areas | 586 (38.2) | 405 (40.7) | 181 (33.8) |
| Mean number of hours per day (SD) | 2.0 (1.7) | 2.1 (1.8) | 1.9 (1.6) |
| Range | 0.5 - 14 | 0.5 - 14 | 0.5 - 12 |
| Drink caffeinated tea | 1043 (68.1) | 668 (67.1) | 375 (69.9) |
| Mean number of cups per day (SD) | 2 (1.4) | 2 (1.3) | 2.3 (1.6) |
| Range | 0.5 - 12 | 0.5 - 8 | 0.5 - 12 |
| Information not provided | 34 (2.2) | 21 (2.1) | 13 (2.4) |
| Drink caffeinated coffee | 1120 (73.1) | 761 (76.5) | 359 (66.9) |
| Mean number of cups per day (SD) | 2 (1.2) | 2.1 (1.3) | 1.8 (1.2) |
| Range | 0.5 - 10 | 0.5 - 10 | 0.5 - 10 |
| Information not provided | 34 (2.2) | 21 (2.1) | 13 (2.4) |
| Drink other caffeinated beverages | 371 (24.2) | 271 (27.2) | 100 (18.7) |
| Average volume in ml per day (SD) | 330.8<br>(336.6) | 335.8<br>(340.2) | 317.3<br>(330.7) |
| Range | 10 - 2000 | 10 - 2000 | 10 - 2000 |
| Information not provided | 35 (2.3) | 22 (2.2) | 13 (2.4) |

**Note:** SD – standard deviation; Sample N=1532

Three per cent of the cohort are current cigarette, cigar, or pipe smokers and smoke on average 13 ± 8 cigarettes/cigars/pipes per day. Thirty-six per cent of the sample) reported being former smokers and disclosed smoking 15 ± 11 cigarettes/cigars/pipes a day in the past. For current smokers, there was a significant negative relationship between the number of cigarettes/cigars/pipes smoked daily and the age at onset of the first symptom of PD (Pearson correlation coefficient r = −0.2, *p* =0.03).

Many participants reported being exposed to second-hand smoke at home (56%), at work (46%) and in other areas (38%), with the mean of hours of passive smoking ranging from 2 to 5 (For details refer to Table 4).

Sixty-eight per cent of participants drink tea, 73% drink coffee, and 24% consume other caffeinated beverages. On average, participants drink 2 cups of caffeinated tea or coffee daily, ranging from half a cup to 12 cups a day. There was a significant weak positive correlation between the number of teacups consumed daily and the age at onset of PD symptoms (Pearson correlation coefficient r = 0.1, *p* = 0.001). Those who reported drinking other caffeinated beverages indicated that they consumed on average 330 ± 335 ml of other caffeinated drinks per day. Sex differences in the reported outcomes related to PD’s lifestyle factors are given in **Table 4**.

### Parkinson’s disease comorbidities

Participants reported on a range of comorbidities. PD co-occurring medical conditions included allergy, cancer, melanoma, psychological, cardiovascular, respiratory, endocrine, and gastrointestinal disorders. Back problems were the most common medical condition in 47% of the respondents, and eczema was the least prevalent disorder reported by 7% of the participants **(Table 5)**.

**Table 5.** Self-reported history of comorbidities in the study sample

| Condition / Disorder | N | % |
| --- | --- | --- |
| Back problems | 722 | 47.1 |
| Constipation | 533 | 36.1 |
| Depression | 514 | 33.6 |
| High blood pressure | 506 | 33.0 |
| High cholesterol | 430 | 28.1 |
| Frequent acid reflux | 377 | 24.6 |
| Cold sores | 336 | 21.9 |
| Migraine | 264 | 17.2 |
| Anxiety | 262 | 17.1 |
| Melanoma | 243 | 15.9 |
| Sleep apnoea | 241 | 15.7 |
| Insomnia | 202 | 13.2 |
| Cancer | 201 | 13.1 |
| Asthma / COPD | 194 | 12.7 |
| Pneumonia | 192 | 12.5 |
| REM Sleep behaviour disorder | 177 | 11.6 |
| Diabetes | 156 | 10.2 |
| Osteoporosis | 126 | 8.2 |
| Eczema | 107 | 7.0 |

**Note:** COPD – chronic obstructive pulmonary disease; Sample N=1,532

## Discussion

The present study describes sample characteristics of 1,532 Australians with PD recruited for the Australian Parkinson’s Genetics Study to unravel environmental and genetic associations with PD, along with PD heterogeneity, anti-PD medication response and adverse drug reactions.

Parkinson’s disease incidence varies with gender, age, and race or ethnicity [38,39]. We observed a higher prevalence of male participants than females (65% of males versus 35% of females), in line with most previous PD studies in various populations worldwide [38–40]. The causes of higher male-to-female ratios in PD are unknown, and it is still unclear whether male sex is an independent risk factor for PD. Although estrogens are believed to be neuroprotective in females [41], further research is needed to elucidate whether males could be at greater PD risk due to higher exposure to environmental or occupational risk factors.

The present study is the first to employ an assisted mailout approach to recruit PD participants from all Australian states. We found that individuals of European descent (92% of the sample group) were the most prevalent in the APGS, with a lower proportion of participants from Asian background (4%), Indigenous Australians and Torres Strait Islanders (1%), Africans (0.1%), Pacific Islanders (0.6%), Hispanics (0.3%), and in individuals of mixed ancestry (1.4%). We cannot estimate differences in PD prevalence across ethnicities, given the lack of unaffected controls. Previous Australian epidemiological studies have reported PD prevalence rates between 66 and 415 per 100,000 Australians [42–44]. Similarly, worldwide PD prevalence has been consistently reported between 60 and 335 per 100,000 individuals, with a broad spectrum of ethnic backgrounds identified. For instance, the Israel population, Native Americans, and Alaska Natives have the highest prevalence (335/100 000 for Jewish Israeli and 355/100 000 for Native Americans and Alaska Natives) [45,46]. At the same time, Asian and African communities report the lowest prevalence rates [47–49]. The significantly increased prevalence of PD among distinct ethnic groups is partly attributable to a higher occurrence of PD-linked mutations in these populations (e.g., *LRRK2* G2019S and *GBA* in *Ashkenazi* Jews) [50,51]. However, other factors such as variation in age groups surveyed, sampling methods, and diagnostic and screening criteria cannot be ruled out.

A Parkinson’s disease diagnosis is rare before 50, yet a significant number of patients present early-stage non-motor PD symptoms in the late fifties [1]. Therefore, aging remains the primary risk factor for developing idiopathic PD [52–54], with the incidence of new cases increasing up to 10-fold between 60 and 90 years of age [38,55,56]. In our study, the mean age of onset for PD symptoms was 56 years, suggesting that self-selected PD participants may be among the younger age group, which tends to be more technology-savvy and motivated to participate in medical research. However, aging alone is insufficient to cause PD, and a complex relationship between aging and genetic and environmental factors prompts the disease onset [57]. Aging-associated impairments [58,59] and degeneration of dopaminergic neurons [60] may explain the steep rise in PD prevalence with age.

Previous studies have shown that a traumatic brain injury with loss of consciousness early in life is associated with late-life neurodegenerative conditions, accumulation of Lewy bodies, PD progression, and increased PD risk [61]. However, the pathogenesis of brain injury and its link with neurodegenerative disorders has not been fully elucidated. We identified a significant positive association between the age of head injury and the age at onset of PD symptoms. Participants who had a head injury early in life also reported an earlier age at which the first PD symptoms were manifested. However, only a small percentage of participants reported a lifetime traumatic head injury. Thus, our results appear to support the hypothesis that traumatic brain injury may initiate or exacerbate PD. This may be due to persistent inflammation, metabolic dysfunction, and abnormal protein aggregation [62].

Lifestyle may affect PD susceptibility. For instance, alcohol [15,63,64], tobacco [65,66] and caffeine [67,68] consumption are negatively correlated with PD risk. Previous studies have found that alcohol users have a lower PD risk than non-alcohol users, and those who drink more are less susceptible to PD than those who drink less [64]. Excessive alcohol consumption has been found to reverse this protective effect, increasing PD susceptibility independent of gender [69]. We observed a positive association between alcohol use and PD age of onset in the APGS sample, whereby participants with PD who had more alcoholic drinks consumed per week reported a later age of onset of PD compared to those who consumed fewer alcoholic drinks weekly. This interim observation agrees with previous findings and may imply that alcohol has a protective effect and delays the onset of PD. However, this needs to be explored further due to the possibility of ascertainment bias (e.g., alcohol consumption has negative effects on health and increases the risk of mortality).

Several longitudinal studies have reported an inverse association between smoking and PD. Compared with never smokers, a lower PD risk was observed among current and past smokers [65,70]. Notably, smoking duration and intensity have been inversely related to PD risk, with PD susceptibility decreasing up to 70% when the duration of smoking in years increased [65]. In contrast, we observed an earlier age of symptoms onset in those who smoked more cigarettes/cigars/pipes per day than those who smoked fewer. This should be interpreted with caution, given the small sample size. Also, a recent Mendelian randomisation study [71] suggested that continuing the smoking habit rather than smoking heaviness (i.e., numbers of cigarettes smoked per day) may exert the protective effect of smoking over PD risk.

Prospective studies assessing lifetime coffee and tea consumption and PD highlight a negative correlation between the caffeine in coffee or tea and PD risk [72], and the effect is more pronounced among men than women who regularly drink coffee or tea [67,73,74]. But there is substantial between-study heterogeneity across reported effect estimates. Our correlations between caffeinated tea consumption and age of onset of PD are similar to those previously reported by others [75], with regular tea consumers reporting a later age of symptom onset than non-habitual tea consumers. The precise mechanisms underlying this relationship between lifestyle variables and PD susceptibility are poorly understood.

Consistent with previous findings, we observed high rates of comorbidities including constipation [76,77], depression [78,79], anxiety disorder [80], melanoma [81,82] and diabetes [83]. In particular, the incidence of melanoma is known to be two-fold higher in patients with PD than among the general population [84–86]. In addition, the Australian population exhibits high rates of melanoma due to predominantly fair-skin populations living under high levels of UV radiation. Further investigation is needed to unravel the underlying mechanisms linking PD with melanoma. Although the prevalence estimates of common comorbidities in our cohort are higher than in the general population [87–90], our results support the body of evidence that these particular conditions are substantially more common among individuals with PD and thus may share a common aetiology.

The study design and data analysis methods limitations warrant some cautions when interpreting our interim findings. Notably, our mailout targeted participants aged 40-75 years old regardless of their disease duration since onset. We acknowledge that a large proportion of PD cases have an onset above 75 [91], and thus, our selected age range could have led to a selection bias as many individuals with PD could have been missing from the study. It would be ideal to recruit and follow up all patients at diagnosis or at least from the early stages of the disease. In future recruitment phases, we will prioritise the recruitment of older participants and employ other recruitment methods such as movement disorder clinics, patient support groups, and a public media campaign.

Furthermore, although the 9% response rate in the pilot mailout is comparatively higher than similar studies targeted at individuals with depression, bipolar disorder, alcohol dependence and Alzheimer’s disease [92], it remains low and susceptible to selection bias. In the next stage of our study, we plan to compare the background characteristics of participants recruited through different recruitment channels to identify differences in response rates and potential selection biases. This will also help us identify the most efficient mode of recruitment.

The current pilot study did not have a comparator age- and sex-matched control cohort. We also hope to recruit unaffected controls via referral from PD participants in the future. We will ask each participant to invite one or two friends of the same sex and within a 10-year age range from similar regional and ethnic backgrounds but without PD.

Although most participants (97%) reported being diagnosed with PD by a neurologist or general practitioner, ideally, participants included in a PD study would have their diagnosis confirmed by case note review or in-person evaluation to validate the identification of PD, but this was not feasible in our pilot phase due to limited research funds and a large number of participants.

In summary, we report the baseline characteristics of the APGS pilot cohort. The APGS aims to characterise and improve our understanding of the sociodemographic, genetic, and environmental basis of PD susceptibility, symptoms, and progression in Australia. We used an innovative and highly efficient recruitment approach through assisted mailouts, establishing feasibility and laying the groundwork for expanding and extending the study’s scale and reach in the future. The results of this study will provide an important opportunity for generating evidence on the epidemiology and genetic aetiology of PD in the country. Ultimately, a more extensive representation of diverse Australian participants in worldwide PD studies may lead to a better understanding of the causes of PD and help in the discovery and validation of novel therapeutic targets and the development of new therapies and interventions to prevent, stop, or modify the clinical course of PD in Australia and the rest of the world.

## Data Availability

Data is available upon reasonable request and ethics review and approval for research purposes, in anonymised form, to preserve participants' identities. Inquiries from potential scientific collaborators can be directed to the corresponding authors

## Acknowledgments

We thank all the participants for kindly giving their time to contribute to this study. We thank the Health Data Analysis and Strategy Branch team at Services Australia for their kind assistance and facilitating the mailout.

## Author’s contributions

MER designed the APGS study with input from GDM, JG, PM, PCP, PAL, DZL, PMV, SEM, CRS and NGM. AM, AIC, BLM, LMGM revised and tested the questionnaire and provided intellectual input into the content. RP coordinated the project and sample collection with help from SC and MF. SV cleaned and analysed the data for this cohort profile manuscript. SV and MER drafted the manuscript. All co-authors revised the article for intellectual content. All authors have read and approved the final version of the manuscript.

## Ethics approval

This study has been approved by the Human Research Ethics Committee of the QIMR Berghofer Medical Research Institute (P3711).

## References

1 Poewe W, Seppi K, Tanner CM, et al. Parkinson disease. Nat Rev Dis Primers 2017;3:17013.

2 Pringsheim T, Jette N, Frolkis A, et al. The prevalence of Parkinson’s disease: a systematic review and meta-analysis. Mov Disord 2014;29:1583–90.

3 Deloitte Access Economics. Living with Parkinson’s Disease. 2016. https://www2.deloitte.com/au/en/pages/economics/articles/living-with-parkinsons-disease.html (accessed 30 Jan 2021).

4 Bohingamu Mudiyanselage S, Watts JJ, Abimanyi-Ochom J, et al. Cost of Living with Parkinson’s Disease over 12 Months in Australia: A Prospective Cohort Study. Parkinsons Dis 2017;2017:5932675.

5 GBD 2016 Parkinson’s Disease Collaborators. Global, regional, and national burden of Parkinson’s disease, 1990-2016: a systematic analysis for the Global Burden of Disease Study 2016. Lancet Neurol 2018;17:939–53.

6 Bellou V, Belbasis L, Tzoulaki I, et al. Environmental risk factors and Parkinson’s disease: An umbrella review of meta-analyses. Parkinsonism Relat Disord 2016;23:1–9.

7 Menegon A, Board PG, Blackburn AC, et al. Parkinson’s disease, pesticides, and glutathione transferase polymorphisms. Lancet 1998;352:1344–6.

8 Dissanayaka NNW, Sellbach A, Matheson S, et al. Anxiety disorders in Parkinson’s disease: prevalence and risk factors. Mov Disord 2010;25:838–45.

9 Noyce AJ, Bestwick JP, Silveira-Moriyama L, et al. Meta-analysis of early nonmotor features and risk factors for Parkinson disease. Ann Neurol 2012;72:893–901.

10 Jafari S, Etminan M, Aminzadeh F, et al. Head injury and risk of Parkinson disease: a systematic review and meta-analysis. Mov Disord 2013;28:1222–9.

11 Pezzoli G, Cereda E. Exposure to pesticides or solvents and risk of Parkinson disease. Neurology 2013;80:2035–41.

12 van der Mark M, Brouwer M, Kromhout H, et al. Is pesticide use related to Parkinson disease? Some clues to heterogeneity in study results. Environ Health Perspect 2012;120:340–7.

13 Domínguez-Baleón C, Ong JS, Scherzer CR. Genetic evidence for protective effects of smoking and drinking behavior on Parkinson’s disease: A Mendelian Randomization study. medRxiv Published Online First: 2020.https://www.medrxiv.org/content/10.1101/2020.04.20.20073247v1.abstract

14 Checkoway H, Nielsen SS, Racette BA. The search for environmental risk factors for Parkinson disease. Current Topics in Occupational Epidemiology. 2013;:31–41. doi:10.1093/med/9780199683901.003.0003

15 Bettiol SS, Rose TC, Hughes CJ, et al. Alcohol Consumption and Parkinson’s Disease Risk: A Review of Recent Findings. J Parkinsons Dis 2015;5:425–42.

16 Przedborski S. The two-century journey of Parkinson disease research. Nat Rev Neurosci 2017;18:251–9.

17 Santiago JA, Bottero V, Potashkin JA. Biological and Clinical Implications of Comorbidities in Parkinson’s Disease. Frontiers in Aging Neuroscience. 2017;9. doi:10.3389/fnagi.2017.00394

18 Poortvliet PC, O’Maley K, Silburn PA, et al. Perspective: Current Pitfalls in the Search for Future Treatments and Prevention of Parkinson’s Disease. Front Neurol 2020;11:686.

19 Wirdefeldt K, Gatz M, Reynolds CA, et al. Heritability of Parkinson disease in Swedish twins: a longitudinal study. Neurobiol Aging 2011;32:1923.e1–8.

20 Hernandez DG, Reed X, Singleton AB. Genetics in Parkinson disease: Mendelian versus non-Mendelian inheritance. J Neurochem 2016;139 Suppl 1:59–74.

21 Billingsley KJ, Bandres-Ciga S, Saez-Atienzar S, et al. Genetic risk factors in Parkinson’s disease. Cell Tissue Res 2018;373:9–20.

22 Nalls MA, Blauwendraat C, Vallerga CL, et al. Identification of novel risk loci, causal insights, and heritable risk for Parkinson’s disease: a meta-analysis of genome-wide association studies. Lancet Neurol 2019;18:1091–102.

23 Hindorff LA, Bonham VL, Brody LC, et al. Prioritizing diversity in human genomics research. Nat Rev Genet 2018;19:175–85.

24 Mehta P, Kifley A, Wang JJ, et al. Population prevalence and incidence of Parkinson’s disease in an Australian community. Intern Med J 2007;37:812–4.

25 Peters CM, Gartner CE, Silburn PA, et al. Prevalence of Parkinson’s disease in metropolitan and rural Queensland: a general practice survey. J Clin Neurosci 2006;13:343–8.

26 Politi C, Ciccacci C, Novelli G, et al. Genetics and Treatment Response in Parkinson’s Disease: An Update on Pharmacogenetic Studies. Neuromolecular Med 2018;20:1–17.

27 Li B-D, Bi Z-Y, Liu J-F, et al. Adverse effects produced by different drugs used in the treatment of Parkinson’s disease: A mixed treatment comparison. CNS Neurosci Ther 2017;23:827–42.

28 Deng H, Wang P, Jankovic J. The genetics of Parkinson disease. Ageing Res Rev 2018;42:72–85.

29 Gao Y, Wilson GR, Salce N, et al. Genetic Analysis of in an Early-Onset Parkinson’s Disease Cohort. Front Neurol 2020;11:523.

30 Bentley SR, Bortnick S, Guella I, et al. Pipeline to gene discovery - Analysing familial Parkinsonism in the Queensland Parkinson’s Project. Parkinsonism Relat Disord 2018;49:34–41.

31 Poortvliet PC, Gluch A, Silburn PA, et al. The Queensland Parkinson’s Project: An Overview of 20 Years of Mortality from Parkinson’s Disease. J Mov Disord 2021;14:34–41.

32 Goetz CG, Fahn S, Martinez-Martin P, et al. Movement Disorder Society-sponsored revision of the Unified Parkinson’s Disease Rating Scale (MDS-UPDRS): Process, format, and clinimetric testing plan. Mov Disord 2007;22:41–7.

33 Pachana NA, Byrne GJ, Siddle H, et al. Development and validation of the Geriatric Anxiety Inventory. Int Psychogeriatr 2007;19:103–14.

34 Yesavage JA, Sheikh JI. 9/Geriatric Depression Scale (GDS). US. 1986.

35 Visser M, Verbaan D, van Rooden SM, et al. Assessment of psychiatric complications in Parkinson’s disease: The SCOPA-PC. Mov Disord 2007;22:2221–8.

36 Martinez-Martin P, Visser M, Rodriguez-Blazquez C, et al. SCOPA-sleep and PDSS: two scales for assessment of sleep disorder in Parkinson’s disease. Mov Disord 2008;23:1681–8.

37 IBM Corp. (Armonk, NY). IBM SPSS Statistics. 2015.

38 Van Den Eeden SK, Tanner CM, Bernstein AL, et al. Incidence of Parkinson’s disease: variation by age, gender, and race/ethnicity. Am J Epidemiol 2003;157:1015–22.

39 Baldereschi M, Di Carlo A, Rocca WA, et al. Parkinson’s disease and parkinsonism in a longitudinal study: two-fold higher incidence in men. ILSA Working Group. Italian Longitudinal Study on Aging. Neurology 2000;55:1358–63.

40 Solla P, Cannas A, Ibba FC, et al. Gender differences in motor and non-motor symptoms among Sardinian patients with Parkinson’s disease. J Neurol Sci 2012;323:33–9.

41 Cereda E, Barichella M, Cassani E, et al. Reproductive factors and clinical features of Parkinson’s disease. Parkinsonism Relat Disord 2013;19:1094–9.

42 Jenkins AC. Epidemiology of parkinsonism in Victoria. Med J Aust 1966;2:496–502.

43 McCann SJ, LeCouteur DG, Green AC, et al. The epidemiology of Parkinson’s disease in an Australian population. Neuroepidemiology 1998;17:310–7.

44 Chan DK, Dunne M, Wong A, et al. Pilot study of prevalence of Parkinson’s disease in Australia. Neuroepidemiology 2001;20:112–7.

45 Chillag-Talmor O, Giladi N, Linn S, et al. Use of a refined drug tracer algorithm to estimate prevalence and incidence of Parkinson’s disease in a large israeli population. J Parkinsons Dis 2011;1:35–47.

46 Gordon PH, Mehal JM, Holman RC, et al. Parkinson’s disease among American Indians and Alaska natives: a nationwide prevalence study. Mov Disord 2012;27:1456–9.

47 Bower JH. Understanding Parkinson disease in sub Saharan Africa: A call to action for the international neurologic community. Parkinsonism Relat. Disord. 2017;41:1–2.

48 McInerney-Leo A, Gwinn-Hardy K, Nussbaum RL. Prevalence of Parkinson’s disease in populations of African ancestry: a review. J Natl Med Assoc 2004;96:974–9.

49 Wang SJ, Fuh JL, Teng EL, et al. A door-to-door survey of Parkinson’s disease in a Chinese population in Kinmen. Arch Neurol 1996;53:66–71.

50 Orr-Urtreger A, Shifrin C, Rozovski U, et al. The LRRK2 G2019S mutation in Ashkenazi Jews with Parkinson disease: is there a gender effect? Neurology 2007;69:1595–602.

51 Sidransky E, Nalls MA, Aasly JO, et al. Multicenter analysis of glucocerebrosidase mutations in Parkinson’s disease. N Engl J Med 2009;361:1651–61.

52 Bennett DA, Beckett LA, Murray AM, et al. Prevalence of parkinsonian signs and associated mortality in a community population of older people. N Engl J Med 1996;334:71–6.

53 Morens DM, Davis JW, Grandinetti A, et al. Epidemiologic observations on Parkinson’s disease: incidence and mortality in a prospective study of middle-aged men. Neurology 1996;46:1044–50.

54 Tanner CM, Goldman SM. Epidemiology of Parkinson’s disease. Neurol Clin 1996;14:317–35.

55 Twelves D, Perkins KSM, Counsell C. Systematic review of incidence studies of Parkinson’s disease. Mov Disord 2003;18:19–31.

56 Savica R, Grossardt BR, Bower JH, et al. Incidence and pathology of synucleinopathies and tauopathies related to parkinsonism. JAMA Neurol 2013;70:859–66.

57 Pang SY-Y, Ho PW-L, Liu H-F, et al. The interplay of aging, genetics and environmental factors in the pathogenesis of Parkinson’s disease. Transl Neurodegener 2019;8:23.

58 Kaushik S, Cuervo AM. Proteostasis and aging. Nat Med 2015;21:1406–15.

59 Collier TJ, Lipton J, Daley BF, et al. Aging-related changes in the nigrostriatal dopamine system and the response to MPTP in nonhuman primates: diminished compensatory mechanisms as a prelude to parkinsonism. Neurobiol Dis 2007;26:56–65.

60 Collier TJ, Kanaan NM, Kordower JH. Aging and Parkinson’s disease: Different sides of the same coin? Mov Disord 2017;32:983–90.

61 Crane PK, Gibbons LE, Dams-O’Connor K, et al. Association of Traumatic Brain Injury With Late-Life Neurodegenerative Conditions and Neuropathologic Findings. JAMA Neurol 2016;73:1062–9.

62 Delic V, Beck KD, Pang KCH, et al. Biological links between traumatic brain injury and Parkinson’s disease. Acta Neuropathol Commun 2020;8:45.

63 Eriksson A-K, Löfving S, Callaghan RC, et al. Alcohol use disorders and risk of Parkinson’s disease: findings from a Swedish national cohort study 1972-2008. BMC Neurol 2013;13:190.

64 Zhang D, Jiang H, Xie J. Alcohol intake and risk of Parkinson’s disease: a meta-analysis of observational studies. Mov Disord 2014;29:819–22.

65 Thacker EL, O’Reilly EJ, Weisskopf MG, et al. Temporal relationship between cigarette smoking and risk of Parkinson disease. Neurology 2007;68:764–8.

66 Gallo V, Vineis P, Cancellieri M, et al. Exploring causality of the association between smoking and Parkinson’s disease. Int J Epidemiol 2019;48:912–25.

67 Ross GW, Abbott RD, Petrovitch H, et al. Association of coffee and caffeine intake with the risk of Parkinson disease. JAMA 2000;283:2674–9.

68 Costa J, Lunet N, Santos C, et al. Caffeine exposure and the risk of Parkinson’s disease: a systematic review and meta-analysis of observational studies. J Alzheimers Dis 2010;20 Suppl 1:S221–38.

69 Eriksson A, Löfving S, Callaghan RC, et al. Alcohol use disorders and risk of Parkinson’s disease: findings from a Swedish national cohort study 1972-2008. Eur J Public Health 2013;23. doi:10.1093/eurpub/ckt126.181

70 Hernán MA, Zhang SM, Rueda-deCastro AM, et al. Cigarette smoking and the incidence of Parkinson’s disease in two prospective studies. Ann Neurol 2001;50:780–6.

71 Domínguez-Baleón C, Ong J-S, Scherzer CR, et al. Understanding the effect of smoking and drinking behavior on Parkinson’s disease risk: a Mendelian randomization study. Sci Rep 2021;11:13980.

72 Hu G, Bidel S, Jousilahti P, et al. Coffee and tea consumption and the risk of Parkinson’s disease. Mov Disord 2007;22:2242–8.

73 Ascherio A, Zhang SM, Hernán MA, et al. Prospective study of caffeine consumption and risk of Parkinson’s disease in men and women. Ann Neurol 2001;50:56–63.

74 Ascherio A, Weisskopf MG, O’Reilly EJ, et al. Coffee consumption, gender, and Parkinson’s disease mortality in the cancer prevention study II cohort: the modifying effects of estrogen. Am J Epidemiol 2004;160:977–84.

75 Kandinov B, Giladi N, Korczyn AD. Smoking and tea consumption delay onset of Parkinson’s disease. Parkinsonism Relat Disord 2009;15:41–6.

76 Cheon S-M, Ha M-S, Park MJ, et al. Nonmotor symptoms of Parkinson’s disease: prevalence and awareness of patients and families. Parkinsonism Relat Disord 2008;14:286–90.

77 Saleem TZ, Higginson IJ, Chaudhuri KR, et al. Symptom prevalence, severity and palliative care needs assessment using the Palliative Outcome Scale: a cross-sectional study of patients with Parkinson’s disease and related neurological conditions. Palliat Med 2013;27:722–31.

78 Aarsland D, Påhlhagen S, Ballard CG, et al. Depression in Parkinson disease--epidemiology, mechanisms and management. Nat Rev Neurol 2011;8:35–47.

79 Schrag A, Barone P, Brown RG, et al. Depression rating scales in Parkinson’s disease: critique and recommendations. Mov Disord 2007;22:1077–92.

80 Richard IH. Anxiety disorders in Parkinson’s disease. Adv Neurol 2005;96:42–55.

81 Dalvin LA, Damento GM, Yawn BP, et al. Parkinson Disease and Melanoma: Confirming and Reexamining an Association. Mayo Clin Proc 2017;92:1070–9.

82 Ferreira JJ, Neutel D, Mestre T, et al. Skin cancer and Parkinson’s disease. Mov Disord 2010;25:139–48.

83 Santiago JA, Potashkin JA. Shared dysregulated pathways lead to Parkinson’s disease and diabetes. Trends Mol Med 2013;19:176–86.

84 Constantinescu R, Elm J, Auinger P, et al. Malignant melanoma in early-treated Parkinson’s disease: the NET-PD trial. Mov Disord 2014;29:263–5.

85 Rugbjerg K, Friis S, Lassen CF, et al. Malignant melanoma, breast cancer and other cancers in patients with Parkinson’s disease. Int J Cancer 2012;131:1904–11.

86 Huang P, Yang X-D, Chen S-D, et al. The association between Parkinson’s disease and melanoma: a systematic review and meta-analysis. Translational Neurodegeneration. 2015;4. doi:10.1186/s40035-015-0044-y

87 World Health Organization. Global Report on Diabetes. World Health Organization 2016.

88 Forootan M, Bagheri N, Darvishi M. Chronic constipation: A review of literature. Medicine 2018;97:e10631.

89 Organization WH, Others. Depression and other common mental disorders: global health estimates. World Health Organization 2017. https://apps.who.int/iris/bitstream/handle/10665/254610/WHO-MSD-MER-2017.2-eng.pdf

90 Ward WH, Farma JM, editors. Cutaneous Melanoma: Etiology and Therapy. Brisbane (AU): : Codon Publications 2018.

91 Macleod AD, Henery R, Nwajiugo PC, et al. Age-related selection bias in Parkinson’s disease research: are we recruiting the right participants? Parkinsonism Relat Disord 2018;55:128–33.

92 Byrne EM, Kirk KM, Medland SE, et al. Cohort profile: the Australian genetics of depression study. BMJ Open 2020;10:e032580.

