## Supplementary Material for "The Australian Parkinson’s Genetics Study (APGS) - pilot (N = 1,532)"

**Supplementary Table 1** Anti-Parkinson’s disease medication name and corresponding PBS code

| **Medication** | **PBS code** | **Dosage or Strength** |
| --- | --- | --- |
| Levodopa + carbidopa | 1242J, 1245M, 1255C, 8970D, 9743T, 9744W | Levodopa (100 - 250 mg) + carbidopa (25 - 50 mg) |
| Levodopa + benserazide | 2225D, 2226E, 2227F, 2228G, 2229H, 2231K, 8218M, 8219N | Levodopa (50 - 200 mg) + benserazide (12.5 - 50 mg) |
| Levodopa + carbidopa + entacapone | 8797B, 8798C, 8799D, 9292C, 9344T, 9345W | Levodopa (50 - 200 mg) + carbidopa (12.5 - 50 mg) + entacapone (200 mg) |
| Amantadine hydrochloride | 3016R | 100 mg |
| Apomorphine hydrochloride hemihydrate | 10950H, 10971K, 11083H, 11093W, 5609F, 5610G, 9607P, 9640J | 10 mg/1 ml injection |
| Cabergoline | 8393R, 8394T | 500 µg – 2 mg |
| Pramipexole dihydrochloride monohydrate | 3418X, 3419Y, 3420B, 3421C, 3422D, 5143Q, 5145T, 9151P, 9152Q, 9153R, 9393J, 9394K | 125 µg – 4 mg |
| Rotigotine | 1140H, 2384L, 2385M, 2410W | 2 – 8 mg/24 hours |
| Benzatropine mesilate | 11249C, 11255J, 11265X, 2362H | 2 mg tablets, 2 mg/mL injection |
| Trihexyphenidyl (benzhexol) hydrochloride | 1109J, 1110K | 2 – 5 mg |
| Rasagiline | 1952R | 1 mg |

**Supplementary Table 2** APGS Invitations and respondents per state or territory of Australia

| **State or Territory** | **Letters sent by state**  **N (%)** | **Respondents by state**  **N (%)** |
| --- | --- | --- |
| New South Wales | 6280 (31.1) | 6160 (30.2) |
| Victoria | 5020 (25.1) | 4440 (22.2) |
| Queensland | 4220 (21.1) | 4860 (24.3) |
| South Australia | 1880 (9.4) | 1880 (9.4) |
| Western Australia | 1740 (8.7) | 1860 (9.3) |
| Tasmania | 640 (3.2) | 460 (2.3) |
| Australian Capital Territory | 220 (1.1) | 340 (1.7) |
| Northern Territory | 0.3 | 0.6 |

**Supplementary Figure 1** Male sample age distribution in APGS

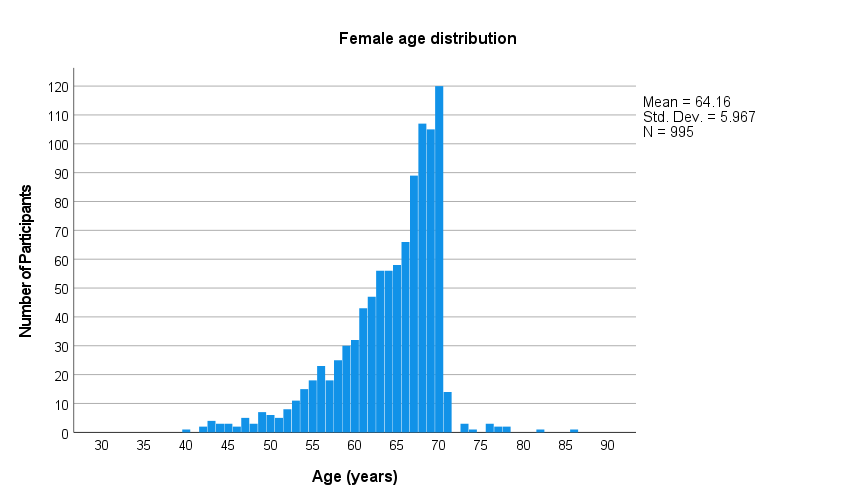

**Supplementary Figure 2** Female sample age distribution in APGS

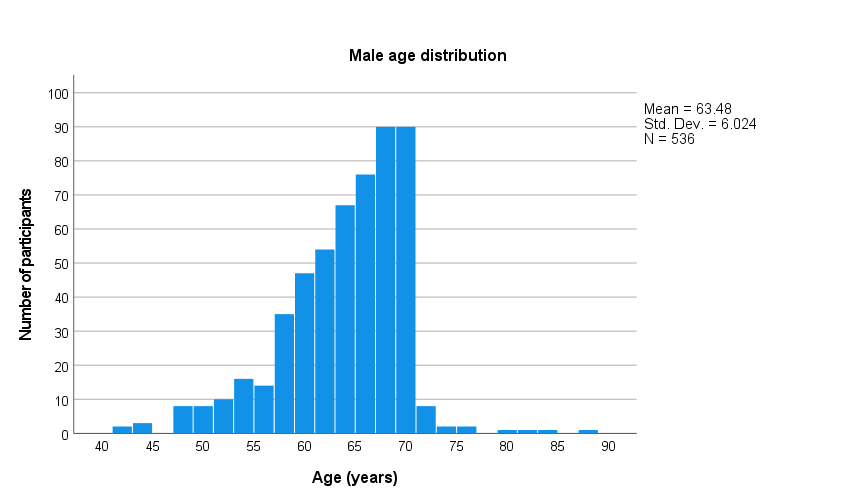
